# Prevalence, Trends, and Associated Factors of Isolated Systolic, Diastolic, and Systolic-Diastolic Hypertension in Peru: A Nine-Year Analysis of the Demographic and Family Health Survey

**DOI:** 10.1101/2024.05.21.24307449

**Authors:** Víctor Juan Vera-Ponce, Fiorella E. Zuzunaga-Montoya, Luisa Erika Milagros Vásquez-Romero, Joan A. Loayza-Castro, Carmen Inés Gutierrez De Carrillo, Enrique Vigil-Ventura

## Abstract

**Introduction:** While HTN is widely seen as a primary threat to cardiovascular conditions worldwide, it is essential to recognize that not all HTN is identical.

**Objective:** To determine the prevalence, trend, and factors associated with each type of HTN: isolated systolic (ISH), isolated diastolic (IDH), and systolic-diastolic (SDH).

**Methods:** A secondary analysis of data from the Demographic and Family Health Survey from 2014– 2022 was conducted. For the analysis of associated factors, a Poisson regression model with robust variance was implemented to calculate adjusted Prevalence Ratios (aPR) along with their 95% confidence intervals.

**Results:** The prevalence was 7.02%, 1.55%, and 3.28% for ISH, IDH, and SDH, respectively. ISH showed a decline in 2022, unlike the other two types, which seem to be on the rise. A statistically significant association was found in men and an increased risk with age for ISH and SDH, unlike IDH, where age acts as a protective factor. Additional factors identified include smoking and excessive alcohol consumption, while a high intake of fruits/vegetables offers a protective effect. Obesity and diabetes were associated with a higher risk, and significant variations by region and altitude, as well as among ethnic groups, were observed.

**Conclusions:** Significant differences in the prevalence of HTN subtypes have been found, underscoring the heterogeneity of this chronic condition, both in related factors and in trends over the years.

## Introduction

Hypertension (HTN), a multifactorial clinical condition typified by persistently elevated arterial blood pressure, is likely to ultimately result in increased susceptibility to developing various long-term complications involving the cardiovascular, cerebrovascular, and renal body systems, among other consequences, if left untreated, according to medical research ^(1)^. The rapidly increasing global ubiquity of HTN presents a formidable test for healthcare frameworks, motivating recent analyses to disentangle the intricacies of HTN and its consequences for public wellness, emphasizing the value of timely administration and preemptive tactics ^(2,3)^.

While HTN is widely seen as a primary threat to cardiovascular conditions worldwide, it is essential to recognize that not all HTN is identical. Current medical literature distinguishes between three main subtypes of HTN: isolated systolic (ISH), isolated diastolic (IDH), and systolic-diastolic (SDH), each with different prevalence profiles across age groups and specific characteristics in the affected individuals ^(4)^. ISH is more common in older adults, while IDH is typically observed in younger adults ^(5)^. However, the condition known as SDH, in which both blood pressure readings are increased, is commonly seen in individuals of middle age and has been linked to an even greater likelihood of developing cardiovascular issues ^(6–9)^.

While the medical literature often treats HTN as a single entity, there is a lack of clear distinction on how its specific subtypes are distributed within the population. This generalization overlooks the crucial differences between the three types. A detailed understanding of the prevalence, trend, and associated factors of every kind of HTN is imperative, as the variability in therapeutic management required for each subtype necessitates a personalized approach ^(10–12)^. Optimizing treatment strategies based on this differentiation is critical to improving health outcomes in the affected population, emphasizing the importance of a more detailed and specific evaluation of HTN in future studies.

## Methods

### Design

An analytical cross-sectional study was conducted through secondary data analysis of the Demographic and Family Health Survey (ENDES) data, provided by the National Institute of Statistics and Informatics (INEI) for the period 2014 to 2022 ^(13)^. This analysis followed the STROBE (Strengthening the Reporting of Observational studies in Epidemiology) guidelines to ensure quality and transparency in reporting observational studies ^(14)^.

### Population, Eligibility Criteria, and Sample

The target population of this study included Peruvian individuals aged between 15 and 99 years, covering both urban and rural areas across all 24 departments of the country. ENDES implemented a probabilistic, stratified, two-stage, and independent sampling design for rural and urban areas. Specific details of the sampling procedure are documented in ENDES technical reports ^(13)^. Individuals aged 18 years or older were selected for analysis, in line with standard definitions of HTN applicable to this age group. Participants not having their blood pressure measured and those reporting a history of HTN were excluded.

### Assessment of Consistency and Plausibility of Measurements

Cutoff thresholds were defined based on methodologies from previous pooled studies to include only blood pressure measurements considered plausible for analysis. Specifically, Systolic Blood Pressure (SBP) measurements were required to be between 70 mmHg and 270 mmHg, and Diastolic Blood Pressure (DBP) measurements were between 50 mmHg and 150 mmHg. Measurements not meeting these plausibility criteria were excluded from the analysis ^(15)^.

### Variables and Measurement

The main variables were as follows ^(4,7,16)^:

- Undiagnosed HTN (which will be mentioned simply as HTN) was defined as those with SBP ≥ 140 mmHg or DBP ≥ 90 mmHg.
- ISH was defined as individuals with SBP ≥ 140 mmHg but with DBP < 90 mmHg, regardless of previous hypertension diagnosis.
- IDH was defined as individuals with DBP ≥ 90 mmHg but with SBP < 140 mmHg.
- SDH was defined as individuals with both SBP ≥ 140 mmHg and DBP ≥ 90 mmHg.

Covariables used were sexo (female and male), Age categorized in years (18 – 35 years, 26 – 59 years, 60 – 69 years, and 70 years and over), Natural Region (Metropolitan Lima, Rest of the coast, Highlands, and Jungle), Educational Level (none, primary, secondary, and higher), Wealth Index (very poor, poor, middle, rich, and very rich), Area of Residence (urban, rural), smoking status (never smoked, former smoker, current smoker, daily smoker), alcohol consumption (never drank or has not drunk in the last 12 months, has not drunk excessively, has drunk excessively), fruit and vegetable intake over 5 servings per day (yes and no), nutritional status measured through body mass index (BMI = Weight (kg)/Height(m)^2^) (normal weight if BMI ≤ 24.99 kg/m^2^, overweight if BMI ≥ 25 to 29.99 kg/m^2^, and obesity ≥ 30 kg/m^2^), Abdominal obesity (if waist circumference ≥ 102 in males or ≥ 88 in females), self-reported diabetes mellitus type 2 (T2DM) (yes, no), Race (Quechua, Aymara, Native or Indigenous Amazonian, Negroid/Black, White/Caucasian, and Mestizo), and Altitude (0 to 499, 500 to 1499, 1500 to 2999, and 3000 or more).

### Procedures

To ensure accuracy and reliability in SBP and DBP measurement, a standardized procedure was adopted using an OMRON brand digital blood pressure monitor, model HEM-713. Two types of cuffs were used to accommodate different arm sizes: one for standard-size arms (220–320 mm) and another for larger arms (330–430 mm).

Blood pressure measurements were taken under controlled conditions, with participants at rest, seated, and with the right arm positioned at heart level on a flat surface. The initial measurement was taken after a five-minute rest period, followed by a second measurement two minutes later, allowing the individual’s cardiovascular conditions to reach a more stable state. The average of these two measurements for SBP and DBP was calculated for each individual, and this average was used for analyses. This method seeks to reduce momentary fluctuations in blood pressure and provides a more accurate and representative measure.

Additionally, ENDES compiled self-reported data from participants on previous diagnoses of HTN, use of antihypertensive medication, and other sociodemographic factors deemed relevant for the study’s analysis.

### Statistical Analysis

In this study, the R 4.03 program will be used for various statistical analyses. Initially, a descriptive analysis of all variables will be conducted, providing an overview of the dataset’s characteristics. Subsequently, to delve deeper into the analysis of associated factors, a Poisson regression model with robust variance will be implemented to calculate adjusted Prevalence Ratios (aPRs), accompanied by their 95% confidence intervals (CI 95%). Finally, for result visualization, various trend graphs covering the period from 2014 to 2022 for each type of HTN will be generated.

### Ethical Considerations

This study used publicly available, anonymized data accessible at no cost to interested parties, ensuring participant privacy by not including information that could personally identify them, thus eliminating potential ethical risks. INEI, in collecting ENDES data, ensured informed consent was obtained from participants before collecting information through the survey.

The database is publicly available for initial consultation and follow-up ^(17)^, offering open access to collected information without personal identifiers and direct contact with the involved human subjects. For these reasons, it was determined that it was not necessary to submit the study for ethics committee evaluation.

## Results

Regarding the general characteristics of the study sample, 51.27% were women, while 13.60% were older adults. A total of 33.62% resided in Metropolitan Lima, the capital of Peru, and 23.75% lived in rural areas. As for lifestyle factors, 11.38% were current smokers, 2.76% consumed alcohol excessively, and only 9.06% consumed at least 5 daily servings of fruits/vegetables. The prevalence based on BMI and WC was 22.76% and 41.69%, respectively. The rest of the characteristics can be seen in Table 1.

**Table 1.**
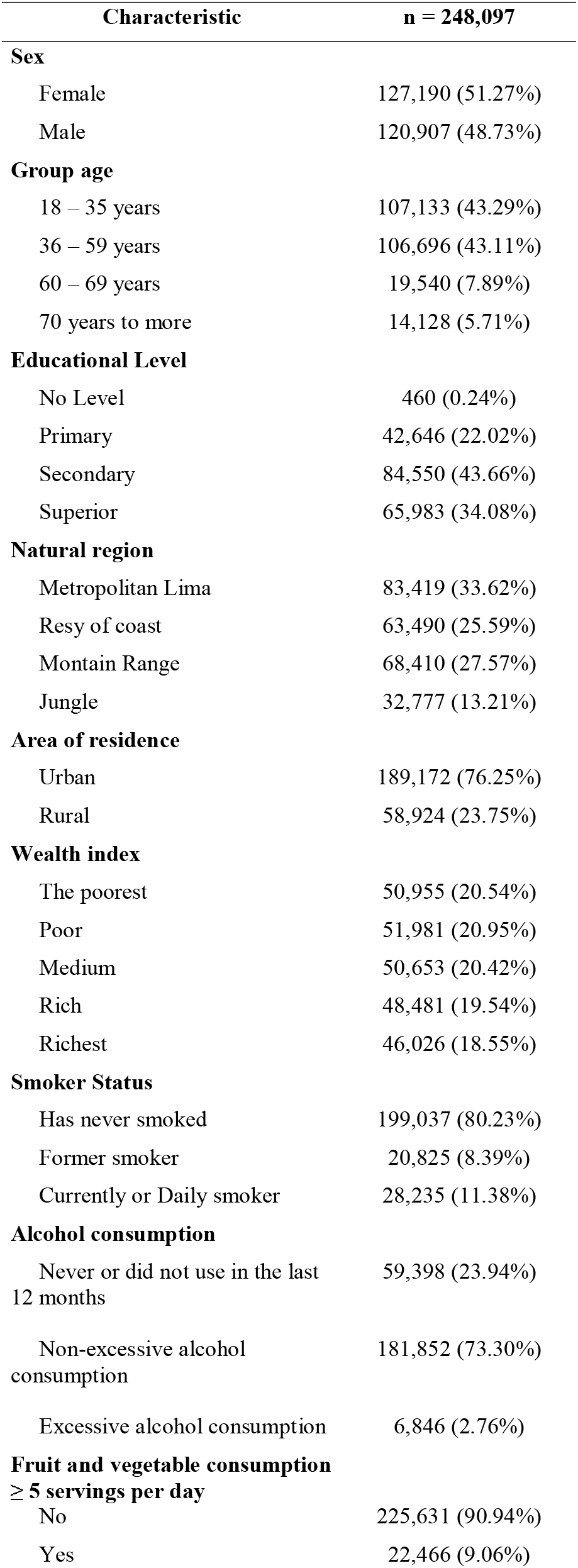
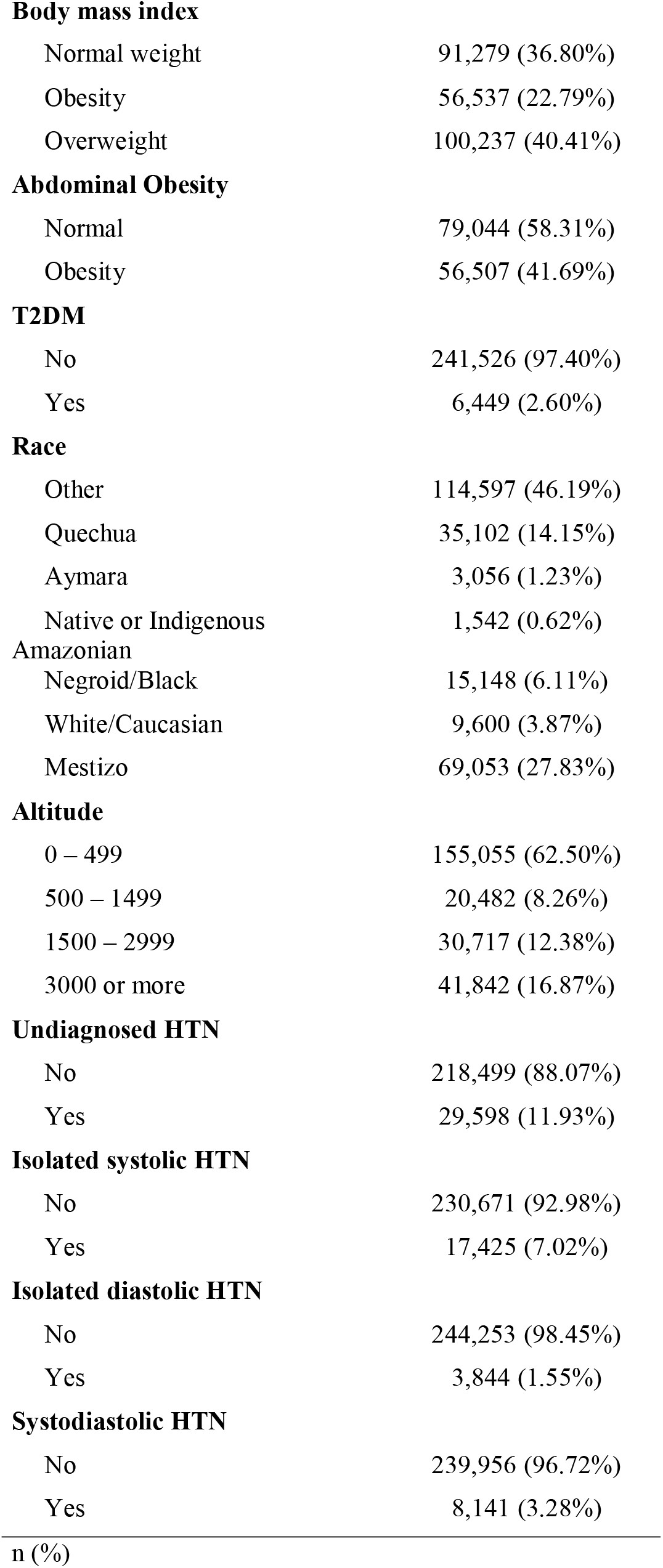
Demographic and anthropometric characteristics of participants.

Regarding the overall prevalence of the types of HTN presented in Figure 1, 11.93% had undiagnosed HTN, while 7.02%, 1.55%, and 3.28% had ISH, IDH, and SDH, respectively. In terms of trends, there has been an increase in the prevalence of undiagnosed HTN over the years, with a slight decrease in 2022; regarding ISH, it showed a decline in the year 2022, unlike the other two, which seems to be increasing.

**Figure 1.**
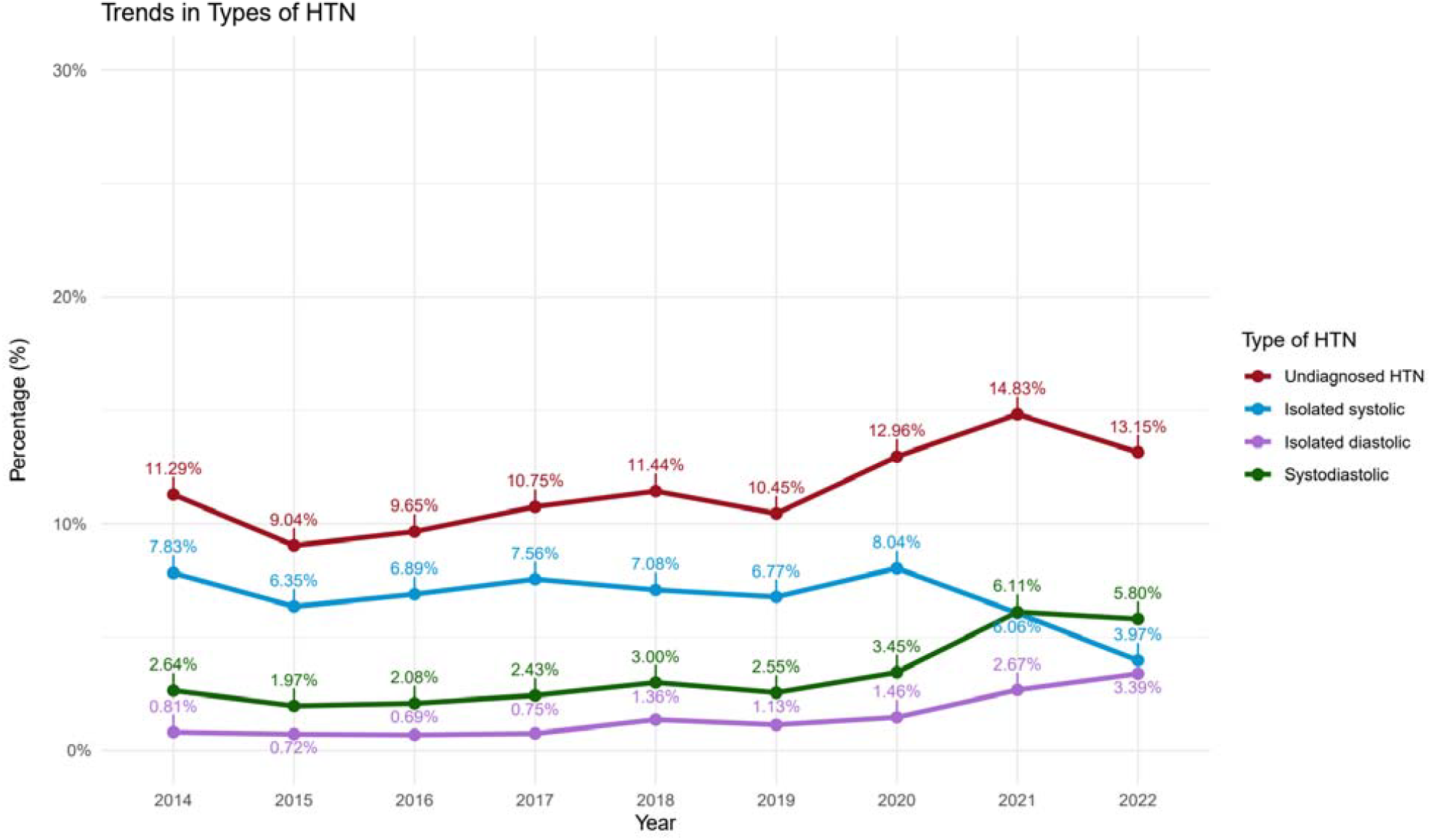
Trend from 2014 to 2022 of the different types of HTN

The multivariable analysis of Table 2 found that males had a higher risk of presenting any HTN, especially SDH. However, regarding age, it was found that older age was associated with a higher risk of undiagnosed HTN, ISH, and SDH; this was not the case for those with IDH, as older age acted as a protective factor. On the other hand, it was found that different regions of the country had a lower probability of presenting any of the types of HTN compared to those living in Metropolitan Lima. The same occurred with living in rural areas. Isolatedly, the wealthy and very wealthy economic classes seemed to have a protective effect against presenting SDH.

**Table 2.** Multivariable regression analysis of the factors associated with the different types of HTN

| Characteristic | Undiagnosed HTN |  | Isolated systolic |  | Isolated diastolic |  | Systodiastolic |  |
| --- | --- | --- | --- | --- | --- | --- | --- | --- |
|  | aPR | 95% CI | aPR | 95% CI | aPR | 95% CI | aPR | 95% CI |
| Sex |  |  |  |  |  |  |  |  |
| Female | — | — | — | — | — | — | — | — |
| Male | 2.53 | 2.43, 2.63 | 2.03 | 1.92, 2.14 | 2.04 | 1.86, 2.24 | 4.11 | 3.82, 4.43 |
| Group age |  |  |  |  |  |  |  |  |
| 18 – 35 years | — | — | — | — | — | — | — | — |
| 36 – 59 years | 2.01 | 1.93, 2.10 | 1.68 | 1.58, 1.79 | 1.53 | 1.41, 1.67 | 3.17 | 2.92, 3.44 |
| 60 – 69 years | 3.62 | 3.43, 3.83 | 4.94 | 4.59, 5.32 | 0.59 | 0.48, 0.71 | 4.47 | 4.02, 4.96 |
| 70 years to more | 4.89 | 4.59, 5.21 | 7.8 | 7.19, 8.45 | 0.27 | 0.17, 0.39 | 3.52 | 3.06, 4.05 |
| Educational Level |  |  |  |  |  |  |  |  |
| No Level | — | — | — | — | — | — | — | — |
| Primary | 0.67 | 0.54, 0.86 | 0.69 | 0.53, 0.91 | 7.85 | 0.82, 11,473 | 0.69 | 0.43, 1.22 |
| Secondary | 0.6 | 0.48, 0.77 | 0.54 | 0.41, 0.71 | 8.38 | 0.88, 12,258 | 0.76 | 0.47, 1.34 |
| Superior | 0.57 | 0.45, 0.73 | 0.47 | 0.36, 0.63 | 9.47 | 0.99, 13,851 | 0.68 | 0.42, 1.20 |
| Natural region |  |  |  |  |  |  |  |  |
| Metropolitan Lima | — | — | — | — | — | — | — | — |
| Resy of coast | 0.72 | 0.69, 0.75 | 0.71 | 0.67, 0.76 | 0.73 | 0.66, 0.80 | 0.71 | 0.67, 0.77 |
| Montain Range | 0.55 | 0.48, 0.62 | 0.52 | 0.43, 0.62 | 0.74 | 0.54, 1.04 | 0.49 | 0.39, 0.62 |
| Jungle | 0.59 | 0.55, 0.63 | 0.58 | 0.52, 0.63 | 0.63 | 0.53, 0.75 | 0.56 | 0.49, 0.63 |
| Area of residence |  |  |  |  |  |  |  |  |
| Urban | — | — | — | — | — | — | — | — |
| Rural | 0.88 | 0.83, 0.95 | 0.92 | 0.84, 1.01 | 0.84 | 0.71, 1.00 | 0.86 | 0.76, 0.97 |
| Wealth index |  |  |  |  |  |  |  |  |
| The poorest | — | — | — | — | — | — | — | — |
| Poor | 0.99 | 0.92, 1.06 | 0.98 | 0.89, 1.08 | 1.16 | 0.97, 1.38 | 0.91 | 0.80, 1.04 |
| Medium | 0.98 | 0.91, 1.05 | 0.99 | 0.90, 1.10 | 1.01 | 0.83, 1.22 | 0.92 | 0.81, 1.06 |
| Rich | 0.95 | 0.88, 1.03 | 0.93 | 0.84, 1.04 | 1.17 | 0.96, 1.42 | 0.85 | 0.74, 0.97 |
| Richest | 0.93 | 0.86, 1.01 | 0.91 | 0.81, 1.02 | 1.28 | 1.05, 1.57 | 0.79 | 0.68, 0.91 |
| Smoker Status |  |  |  |  |  |  |  |  |
| Has never smoked | — | — | — | — | — | — | — | — |
| Former smoker | 0.93 | 0.87, 1.00 | 0.9 | 0.81, 1.00 | 0.96 | 0.84, 1.09 | 0.94 | 0.85, 1.03 |
| Currently or Daily smoker | 1.06 | 1.01, 1.11 | 1.14 | 1.06, 1.22 | 1.14 | 1.02, 1.27 | 0.91 | 0.83, 1.00 |
| Alcohol consumption |  |  |  |  |  |  |  |  |
| Never or did not use in the last 12 months | — | — | — | — | — | — | — | — |
| Non-excessive alcohol consumption | 0.97 | 0.94, 1.01 | 0.94 | 0.80, 1.00 | 1.09 | 0.98, 1.21 | 1 | 0.94, 1.08 |
| Excessive alcohol consumption | 1.08 | 0.99, 1.17 | 0.95 | 0.83, 1.08 | 1.32 | 1.09, 1.60 | 1.17 | 1.01, 1.35 |
| Fruit and vegetable |  |  |  |  |  |  |  |  |
| **consumption ≥ 5 servings per day** |  |  |  |  |  |  |  |  |
| No | — | — | — | — | — | — | — | — |
| Yes | 1.02 | 0.97, 1.07 | 1.06 | 0.98, 1.14 | **0.78** | **0.67, 0.89** | 1.11 | 1.00, 1.24 |
| **Nutricional status** |  |  |  |  |  |  |  |  |
| Normal weight | — | — | — | — | — | — | — | — |
| Obesity | **2.37** | **2.23, 2.51** | **1.72** | **1.59, 1.87** | **4.03** | **3.48, 4.67** | **3.3** | **2.95, 3.69** |
| Overweight | **1.62** | **1.55, 1.70** | **1.43** | **1.34, 1.52** | **2.17** | **1.92, 2.46** | **1.99** | **1.81, 2.19** |
| **Abdominal Obesity** |  |  |  |  |  |  |  |  |
| Normal | — | — | — | — | — | — | — | — |
| Obesity | **1.24** | **1.19, 1.30** | **1.14** | **1.07, 1.22** | **1.07** | **0.96, 1.19** | **1.49** | **1.37, 1.62** |
| **T2DM** |  |  |  |  |  |  |  |  |
| No | — | — | — | — | — | — | — | — |
| Yes | **1.13** | **1.06, 1.21** | **1.18** | **1.07, 1.29** | 1.2 | 0.98, 1.44 | 1.06 | 0.93, 1.20 |
| **Race** |  |  |  |  |  |  |  |  |
| Other | — | — | — | — | — | — | — | — |
| Quechua | **0.92** | **0.86, 0.99** | 1.08 | 0.98, 1.18 | 0.91 | 0.77, 1.09 | **0.73** | **0.64, 0.82** |
| Aymara | 0.9 | 0.79, 1.02 | 1.16 | 0.98, 1.37 | **0.68** | **0.48, 0.94** | **0.67** | **0.52, 0.84** |
| Native or Indigenous Amazonian | 1.12 | 0.92, 1.35 | 1.08 | 0.80, 1.43 | 1.28 | 0.82, 1.93 | 1.11 | 0.78, 1.54 |
| Negroid/Black | 0.99 | 0.92, 1.07 | **1.12** | **1.01, 1.24** | 0.92 | 0.76, 1.11 | 0.88 | 0.77, 1.01 |
| White/Caucasian | 1.03 | 0.95, 1.11 | 0.96 | 0.85, 1.08 | 1 | 0.82, 1.22 | 1.13 | 0.98, 1.29 |
| Mestizo | 0.97 | 0.91, 1.03 | 1.07 | 0.98, 1.17 | 0.92 | 0.79, 1.08 | 0.86 | 0.77, 0.96 |
| **Altitude** |  |  |  |  |  |  |  |  |
| 0 – 499 | — | — | — | — | — | — | — | — |
| 500 – 1499 | 0.94 | 0.88, 1.01 | 1.02 | 0.93, 1.12 | 0.98 | 0.83, 1.16 | **0.79** | **0.69, 0.90** |
| 1500 – 2999 | **1.35** | **1.20, 1.52** | **1.19** | **1.01, 1.41** | **1.44** | **1.05, 1.93** | **1.64** | **1.33, 2.01** |
| 3000 or more | **1.2** | **1.05, 1.38** | 0.98 | 0.80, 1.19 | **1.56** | **1.10, 2.17** | **1.44** | **1.13, 1.83** |

\*each factor has been independently adjusted for sex, group age, natural region, area of residence, education, wealth index, smoker status, alcohol consumption, nutritional status, obesity abdominal, T2DM, consumption of fruits and vegetables, race and altitude,
aRP: Adjusted Prevalence ratio. 95% CI: 95% confidence interval
Source: self made

Current smokers had a higher risk of presenting any HTN, except for SDH. Those who consumed alcohol excessively were more likely to present IDH and SDH. Those consuming more than five portions of fruits/vegetables daily were less likely to present IDH. Regarding obesity by BMI or WC, there was a higher probability of presenting any HTN. Moreover, patients with T2DM were more likely to have undiagnosed HTN and ISH.

Regarding race, Quechuas and Aymaras were less likely to present undiagnosed HTN, IDH, and SDH. Meanwhile, those of Black race were more likely to have ISH. Finally, higher altitude was generally associated with a higher probability of presenting some HTN.

## Discussion

The observed shifts in HTN subtype trends may suggest significant changes in public health program trends. The decline in ISH prevalence in 2022 might result from targeted interventions for older populations, such as exercise programs and healthy diets, effectively reducing SBP ^(18,19)^. This could indicate the partial success of these interventions in reducing the prevalence of this HTN type.

The increase in the prevalence of IDH and SDH could be related to lifestyle changes, such as increased obesity and sedentary behavior, particularly among younger populations. Recent research suggests these risk factors are increasingly prevalent in the general population, potentially explaining the rise in these forms of HTN ^(10,19)^. Furthermore, IDH, previously considered less common and of lower risk, is gaining recognition for its association with long-term cardiovascular risks, especially in young and middle-aged adults ^(9,20)^.

The results suggest that HTN determinants are not uniform across its subtypes. The greater predisposition of men to develop any HTN, particularly SDH, aligns with existing literature indicating gender differences in cardiovascular physiology and HTN risk factors ^(3,21,22)^. However, discrepancies exist regarding other studies that suggest women exhibit a more pronounced elevation in blood pressure ^(10,23)^. Conversely, the relationship between age and HTN type highlights interesting patterns;. In contrast, advanced age is associated with a higher risk of ISH and SDH, a protective effect was observed in IDH. The pathophysiological mechanisms behind this may be aging-related arterial stiffness and vascular elasticity loss contributing to increased systolic pressure, while diastolic pressure may decrease or stabilize ^(22,24,25)^.

The analysis also revealed that living outside Metropolitan Lima and in rural areas is associated with a lower likelihood of presenting any HTN. This finding could reflect differences in lifestyles, exposure to environmental factors, and access to health services between urban and rural areas. A study by Miranda et al. highlights how urbanization and associated lifestyles can increase HTN risk ^(26)^.

The observation that smoking increases the risk of all HTN types, except for SDH, suggests a complex relationship between smoking and blood pressure dynamics. Unlike previous studies that showed a direct relationship between smoking and an increased risk of HTN in general, our results indicate that the impact of smoking may vary by HTN type. This could be due to tobacco’s differentiated effects on vascular function and arterial stiffness, necessitating further research to fully understand its role in SDH. ^(27,28)^.

The link between excessive alcohol consumption and a higher risk of IDH and SDH highlights alcohol’s role in disrupting blood pressure regulation. Alcohol is known to affect the cardiovascular system, including endothelial function alterations and vascular tone regulation, which could explain its association with these specific HTN types. Research into reducing alcohol intake corresponds with claims that it may prove advantageous towards both preventing and handling HTN ^(29,30)^.

The protective impacts of a diet abundant in fruits and vegetables against IDH may be due to the advantages of consuming foods packed with essential nutrients, antioxidants, and fiber which effectively support cardiovascular well-being. Research has revealed that eating more fruits and vegetables in one’s diet is linked to decreased blood pressure readings, potentially as a result of beneficial modifications to how blood vessels work and decreased rigidity in the arteries. This may also indicate that consuming these can specifically protect against certain types of HTN ^(31,32)^.

The association of obesity, measured by both BMI and WC, with an increased risk of all HTN types, underscores obesity as a critical risk factor for HTN. Through various mechanisms such as insulin resistance, systemic inflammation, and alterations in renal and endothelial function that collectively emphasize the significance of weight management for HTN prevention, obesity has been shown to contribute to the onset of HTN ^(28,33)^.

The higher likelihood of having ISH in patients with T2DM highlights the interconnection between both pathologies. Diabetes can exacerbate HTN risk through insulin resistance, endothelial dysfunction, and the accumulation of advanced glycation end-products, reinforcing the need for integrated management of both conditions, specifically affecting endothelial stiffness and increasing myocardial contraction force, leading specifically to an increase in SBP ^(34,35)^.

The findings regarding ethnic differences in HTN prevalence underscore the importance of considering genetic and cultural diversity in HTN study and management. The finding that individuals of Quechua and Aymara origin are less likely to suffer from IDH and SDH suggests that genetic factors or lifestyles associated with these communities may confer a degree of protection against these forms of HTN. Conversely, the higher prevalence of ISH among Black individuals highlights the need for public health strategies that address ethnic disparities in HTN risk ^(36,37)^. This finding emphasizes the importance of implementing personalized and culturally sensitive public health approaches that not only consider general recommendations for HTN prevention and management but also tailor to the specific needs of different ethnic groups. Promoting further research to explore in depth the genetic bases and lifestyle factors contributing to these ethnic differences in HTN prevalence may facilitate the development of more effective and targeted interventions, thus contributing to health equity.

While demographic, socioeconomic, and geographical elements hold significance for HTN prevention and management, these findings especially underscore the necessity of encouraging a lifestyle emphasizing smoking cessation, moderate alcohol intake, a diet rich in fruits and vegetables, weight maintenance, and appropriate diabetes care to promote wellness. Public health interventions aiming to reduce the burden of all HTN types would be most effective if they prioritized lifestyle modifications tailored specifically to the varying needs and risk profiles of diverse population groups.

### Contribution in Public Health

This research reflects the underlying complexity in the epidemiology of HTN, emphasizing the importance of distinguishing between different types of hypertension for accurate diagnosis and appropriate therapeutic management. This study highlights the necessity of tailoring screening and preventative tactics to account for the idiosyncrasies uncovered in the prevalence patterns of distinct HTNs, with personalized strategies considering demographic particulars like age and gender alongside lifestyle habits encompassing the usage of tobacco and alcohol and dietary selections.

Thus, by observing a lower likelihood of presenting HTN in regions outside of Metropolitan Lima and in rural areas, as well as the protective effect of a higher economic position on IDH, the manuscript highlights the importance of addressing socioeconomic disparities and access to health services in public health strategies to combat HTN. Meanwhile, the association found between lifestyle, including tobacco and alcohol consumption, obesity, and T2DM, with the prevalence of different types of HTN underscores the importance of promoting healthy living habits as a central pillar in hypertension prevention.

Through illuminating the tendency to underestimate and underdiagnose high blood pressure within broader society, especially where it remains unseen, the report makes a pressing plea to better develop early discovery tactics and entry to healthcare services. By both enabling early interventions to diminish the frequency of high blood pressure while also helping to curb potential long-term issues linked to poorly regulated hypertension, such as cardiovascular illnesses, kidney deterioration, and brain attacks, this method could serve to dually curb prevalence and complications.

This manuscript makes an exceptionally meaningful addition to the current understanding of health technology assessment and its governance in matters concerning public health. This provides fertile ground for further exploration and the crafting of more discerning and impactful healthcare directives going forward. By determining which risk factors and how prevalence may change for various hypertensive artery types, public health officials can design interventions focused on where needs are greatest, allowing efforts to make a society increasingly robust against this long-term illness by lessening its widespread influence.

### Limitations

This study has several limitations. First, the cross-sectional design prevents us from establishing causality between the identified factors and HTN subtypes. Second, there’s potential bias due to self-reported data, particularly concerning lifestyle factors. Third, while information on certain possible skewing factors, like dietary sodium consumption and exercise habits, was absent which could have impacted outcomes, the lack of data on some potential confounders may have influenced the results.

## Conclusion

This research offers meaningful understanding of the frequency and influencing factors of distinctive high blood pressure classifications within a Peruvian community. The findings underscore the necessity of carefully crafted public health initiatives considering diverse demographic segments’ unique needs and vulnerabilities. Addressing lifestyle factors, promoting healthy behaviors, and considering demographic and socio-economic disparities are crucial steps in reducing the burden of HTN. Further investigation is required to examine the causal links discovered and craft more impactful approaches for avoidance and treatment.

## Data Availability

The data supporting the findings of this study can be accessed in the follow link: https://proyectos.inei.gob.pe/microdatos/

https://proyectos.inei.gob.pe/microdatos/

## Acknowledgments

A special thanks to the members of Instituto de Investigación de Enfermedades Tropicales, Universidad Nacional Toribio Rodríguez de Mendoza de Amazonas (UNTRM), Amazonas, Perú who provided valuable comments during the preparation of this study.

## Informed consent

It was not necessary to obtain informed consent in this Study

## Author contributions

Víctor Juan Vera-Ponce: Conceptualization, Investigation, Methodology, Resources, Writing - Original Draft, Writing - Review & Editing

Fiorella E. Zuzunaga-Montoya: Investigation, Project administration, Writing - Original Draft, Writing - Review & Editing

Luisa Erika Milagros Vásquez-Romero: Investigation, Resources, Writing - Original Draft, Writing - Review & Editing

Joan A. Loayza-Castro: Software, Data Curation, Formal analysis, Writing - Review & Editing

Carmen Inés Gutierrez De Carrillo **r:** Validation, Visualization, Writing - Original Draft, Writing - Review & Editing

Enrique Vigil-Ventura: Methodology, Supervision, Funding acquisition, Writing - Review & Editing

